# The Lupus Epigenome Relates to Genetics, Transcription and Serological Profiles with Dependency on Molecular Subtypes and Informs Drug Discovery

**DOI:** 10.1101/2023.01.19.22283772

**Authors:** Olivia Castellini-Pérez, Guillermo Barturen, Manuel Martínez-Bueno, Andrii Iakovliev, Martin Kerick, Raúl López-Domínguez, Concepción Marañón, Javier Martín, Esteban Ballestar, PRECISEADS Clinical Consortium, PRECISEADS Flow Cytometry Study Group, María Orietta Borghi, Weiliang Qiu, Cheng Zhu, Srinivas Shankara, Athina Spiliopoulou, Emanuele de Rinaldis, Elena Carnero-Montoro, Marta E. Alarcón-Riquelme

## Abstract

**Objective:** The heterogeneity of systemic lupus erythematosus (SLE) can be explained by epigenetic alterations that disrupt transcriptional programs mediating environmental and genetic risk. This study evaluated the epigenetic contribution to SLE heterogeneity considering molecular and serological subtypes, genetics and transcriptional status, followed by drug target discovery.

**Methods:** We performed a stratified epigenome-wide association studies of whole blood DNA methylation from 213 SLE patients and 221 controls. Methylation quantitative trait loci analyses, cytokine and transcription factor activity - epigenetic associations and methylation-expression correlations were conducted. New drug targets were searched for based on differentially methylated genes.

**Results:** In a stratified approach, a total of 974 differential methylation CpG sites with dependency on molecular subtypes and autoantibody profiles were found. Mediation analyses suggested that SLE-associated SNPs in the HLA region exert their risk through DNA methylation changes. Novel genetic variants regulating DNAm in disease or in specific molecular contexts were identified. The epigenetic landscapes showed strong association with transcription factor activity and cytokine levels, conditioned by the molecular context. Epigenetic signals were enriched in known and novel potential drug targets for SLE.

**Conclusion:** This study expands the number of genes associated with SLE and reveals novel pathways of disease. The findings reveal possible genetic drivers and consequences of epigenetic variability on SLE heterogeneity and disentangles the DNAm mediation role on SLE genetic risk and the genetic architecture of DNAm in different molecular contexts. Finally, novel targets for drug development were discovered.

## INTRODUCTION

Systemic lupus erythematosus (SLE) is an autoimmune disease (SAD) caused by the activation of autoreactive T and B cells, the release of inflammatory cytokines and the formation of immune complexes that deposit in tissues, resulting in organ damage. It predominantly appears in young to middle-aged women with a 9:1 female:male bias (1). Treating and diagnosing SLE is challenged by the patients’ heterogeneity in terms of diversity of symptoms, manifestations (2), the organs affected, and a diverse array of autoantibody (AAb) specificities. Although AAb production helps in the diagnosis of some autoimmune diseases and is related to several clinical manifestations, the complexity of SLE is such that patients can present a wide range of specificities, being, moreover, not solely identified in SLE or in specific SLE manifestations.

The understanding of genetic, environmental and molecular mechanisms disrupting immunity and triggering autoantibody production is not well understood. Genome-wide association studies (GWAS) identified several susceptibility genes among which HLA class II locus and genes such as *TNIP, BANK1*, and *IRF5* exhibit the strongest risk effects (3,4). A strong *HLA* genetic association with the presence of autoantibodies such as anti-SSB, anti-SSA, anti-RNP, anti-SM (5) and anti-dsDNA production has been recognized since long (5,6). The most important molecular characteristic of SLE is the overexpression of several interferon-regulated genes (IRG) known as the interferon (IFN) signature, also widely observed at the epigenetic level (epigenIFNsig) in all blood cell types and tissues(7) as well as in other SADS(8,9). A strong interaction between HLA genetic variation, the production of anti-SSA AAb and the epigenetic IFN signature has been reported in primary Sjogren’s syndrome (9), however the genetic and autoantibody determinants of the IFN signature in SLE are still not clear. Recently, the study by *Barturen et al*. (8) molecularly reclassified SLE and other six SADs into different molecular subtypes with important clinical implications. SLE and SADs patients could be stratified into an inflammatory subtype, with increased activity of genes related to the function of monocyte and neutrophil; a lymphoid subtype, with genes related to the function of these immune cells; and an IFN subtype, defined by enhanced activity in genes induced by IFN. This molecular classification needs to be taking into account to overcome SLE heterogeneity.

Heritability studies showed that genetics explains only a small fraction, less than 10% of SLE susceptibility (10–12), suggesting the important contribution of environmental or non-genetic factors (13) and possibly the role of epigenetics mediating gene-by-environment interactions. Epigenetic modifications allow to retrieve different phenotypes from a unique genotype, and are fundamental for immune cells to exhibit diverse and plastic functions responding to evolving environments, stimuli and differentiation processes (14). Dysregulation at the epigenetic level has been identified in association with SLE and some SLE manifestations (15–17). The genome of SLE patients is globally hypomethylated -unmethylated state of CpGs in a normally methylated sequence - and a large percentage of SLE patients exhibit the epigenIFNsig but, intriguingly, not all of them. Functional genomics deciphers the regulatory role of many disease-associated non-coding genetic variants (18), but studies are scarce in SLE (17). Approaches such as methylation quantitative trait loci (meQTL) (18)allows identifying genetic variants influencing disease by modifying DNA methylation (DNAm) (17). However, large meQTL studies have been performed in non-disease populations despite the increasing recognition that regulatory genetic effects can be dependent on age, context and pathological status (9,18,19)

Despite the increasing number of molecular and epigenetic studies in SLE, previous work has not paid attention to disease heterogeneity nor to the possible role of genetic factors or their consequences at the transcriptional and cytokines levels.

The present work moves a step forward in the understanding of epigenetic landscapes in SLE, and their possible drivers and consequences, as well as in the identification of potential new SLE drug targets. We speculate that stratifying EWAS based on molecular subtypes and autoantibody specificities, and integrating different multi-omics layers, provides greater statistical power to find new and group-dependent epigenetic associations that might have a specific genetic regulation and context-specific epigenetic correlation with transcriptional factors and cytokine expression.

## METHODS

An overview of the study design is depicted in **Figure 1**.

**Figure 1.**
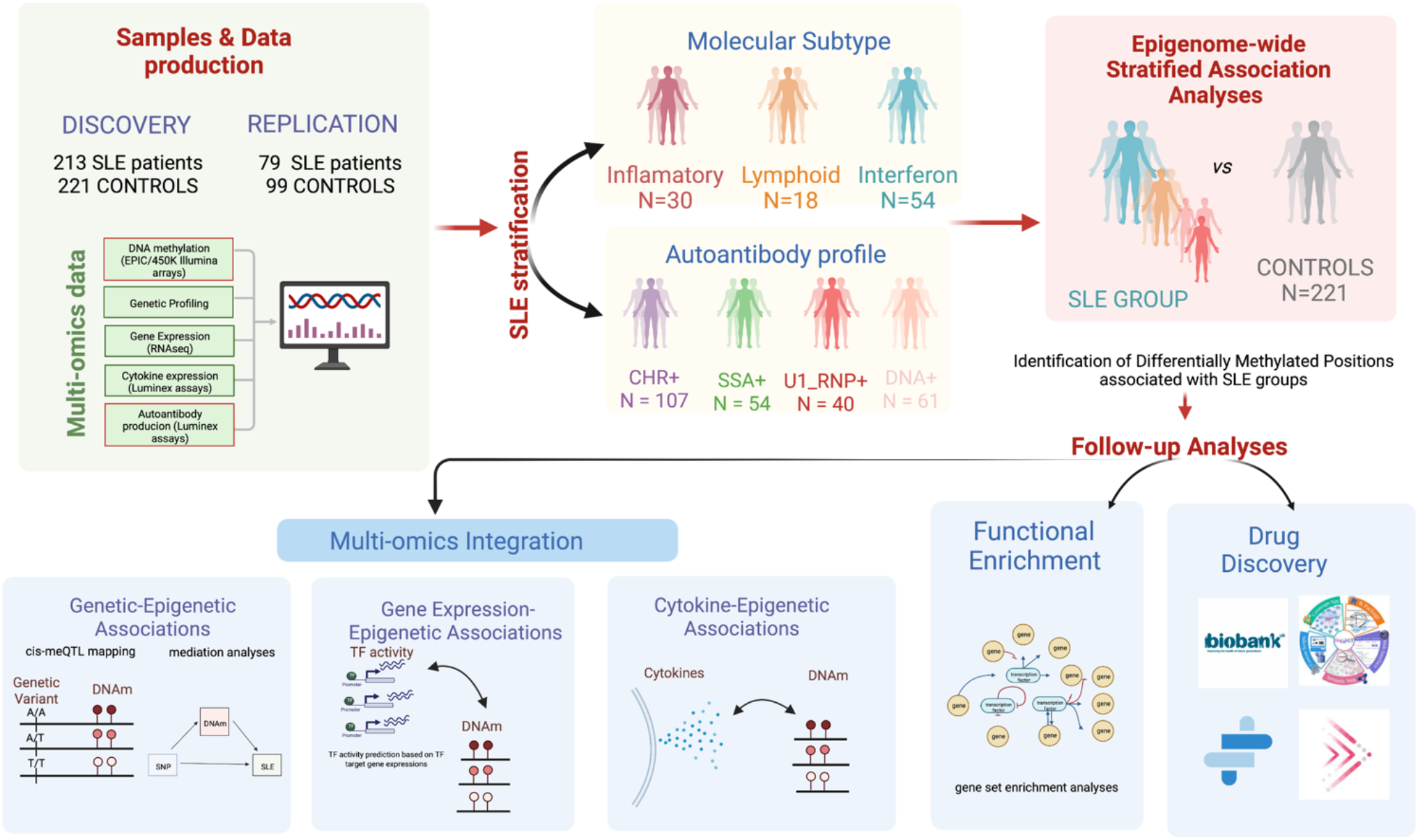
Overview of the Study Design. Schematic outline illustrating the data analysis and the use of patients’ genetic and molecular information to stratify patients into molecularly homogenous groups and autoantibody-positivity profiles to further perform EWAS functional enrichment analyses, meQTL analyses, cytokine association, and drug discovery according to molecular and serological subtypes.

### Samples

We included 292 SLE patients and 320 healthy controls (CTRL). The sample was divided into a discovery set (213 SLE patients and 221 healthy controls with DNAm data based on the EPIC array), and a replication set (79 SLE patients and 99 with DNAm data based on the 450K array) from the PRECISESADS project (8). **Supplementary Table 1** describes the main characteristics of the study sample, together with the groups and traits analyzed. Blood cell proportions were obtained using flow cytometry (20) while autoantibodies, cytokines and other inflammatory mediators were analyzed from serum. Autoantibodies (anti-chromatin, anti-dsDNA, anti-U1RNP, anti-SSA/Ro, anti-SSB/La, anti-SM, anti-β2 glycoprotein 1, anti-β2 microglobulin, IgG anti-cardiolipin, IgM anti-cardiolipin, rheumatoid factor, and anti-ENA), cytokines and inflammatory mediators (BAFF, BLC, CRP, FASL, GDF15, IL1RII, IL1RA, IL6 IP10, MCP2, MCP4, MIP1B, MMP2, MMP8, TARC, TGFβ1, TNFRI and TNFα) were measured in serum samples as described by *Barturen et al*. (8).

### DNA methylation (DNAm) profiling

DNAm data was produced using the Illumina Infinium HumanMethylationEPIC BeadChip array and the Illumina Infinium HumanMethylation450 BeadChip array, which covers up to 850,000 and 485,512 CpG sites respectively. DNA was extracted from peripheral blood samples from which the genome was amplified and hybridized to the Illumina arrays. Standard methodological procedures for quality control and probe filtering were performed as described previously (21). Samples were excluded based on the detection P criteria > 99%, poor bisulfite conversion based on control dashboard check, and sex mismatches according to failed chromosome X and Y clustering. Probes were filtered out based on detection *P* > 0.01 in > 95% of samples. Additionally, probes located at the X and Y chromosomes were separated in different datasets to avoid gender bias. Probes with genetic variants at their CpG sites were also excluded. After applying these filtering steps we obtained 776,284 and 433,337 autosomic probes in the discovery dataset and in the replication dataset, respectively. After QC, the raw methylation beta values were background corrected and normalized using the functional normalization within the meffil R-package. A beta value ranging from 0 to 1 was used to measure DNAm in whole blood, being 1 the methylated status with 100% of cells being methylated at a given CpG and 0 the unmethylated status with 0% of cells being methylated.

### Genetic profiling

Genotyping was performed using InfiniumCore from Illumina. Imputation was performed using the Michigan Imputation Server(22) and Haplotype Reference Consortium as reference panel(23). Filtering of genetic variants and quality controls were performed by PLINK (24). A total of 4,553,097 variants with a minor allele frequency (MAF) higher than 0.05 were used for subsequent analyses.

### Epigenome-wide association analyses (EWAS)

We performed a series of EWAS interrogating DNAm differences between groups at each autosomic CpG site. We used linear regression models adjusting by sex, age and blood cell composition (B cells, CD4 T cells, CD8 T cells, monocytes, neutrophils and natural killer cells) as well as technical confounder effects (Sample_Plate and Sample_Position). SLE-associated differentially methylated CpG positions (SLE-DMPs were identified comparing DNAm between SLE patients and CTRLs. We stratified SLE patients into the molecular subtypes described in *Barturen et al*. (8) and by autoantibody (AAbs) profile and compared their DNAm separately with CTRL. In total, 30 SLE patients were classified into the inflammatory subtype, 18 into the lymphoid subtype and 54 to the IFN subtype in the discovery cohort (**Figure 1**) and 11, 7 and 19 SLE patients in the replication cohort as previously defined by Barturen et al. (8). We only performed EWAS on those AAbs that exhibited positivity in more than 20 SLE patients and corrected models for the presence of all other autoantibodies. We applied a Bonferroni significance threshold (*P* < 5 × 10^−08^) to claim genome-wide significance. We assessed replication (P < 0.05) in an independent cohort from based on 450K-methylation data and determined replication rate for those CpGs present in both arrays. To determine group-dependent DMPs (ex. SLE-dependent DMPs), we used strict significant threshold for the specific DMP passing a Bonferroni threshold in one group while exhibiting a *P* > 0.001 in the others.

### Functional enrichment analyses

Gene-set enrichment function analyses were performed to reveal if genes annotated to DMP were enriched in particular functional pathways. We used the Reactome database (25) implemented by *enrichPathway function* in R library ReactomePA (26). As background distribution we used all genes interrogated in the EPIC array. We separately analyzed DMP with hypomethylation and hypermethylation events.

### Methylation quantitative trait *loci* analysis

We performed methylation quantitative trait locus (meQTL) mapping using the Matrix eQTL R package (27), by means of linear regressions in which the minor allele dosage effect on DNAm was tested while adjusting for age, sex, batch effects, cells proportions, the first genetic principal component, and disease status. We defined *cis*-meQTLs as SNPs located no farther than 1Mb from the interrogated DMP. *Cis*-meQTLs were separately discovered in different groups: i) whole sample, ii) SLE patients, iii) healthy controls, iv) inflammatory SLE patients, and vi) IFN SLE patients. Lymphoid subtype was discarded from this analysis due to the low sample size. We used a permutation based False Discovery Rate (FDR) < 0.05 to claim significance. We investigated the interaction effect between SNP and group for the following groups: SLE vs CTRL and inflammatory vs IFN. To determine group-dependent meQTLs we used the following statistical conditions: FDR < 0.05 in one group and *P* > 0.05 in the other group and *P* < 0.05 significant interaction effect. We also looked for meQTLs with an opposite and significant genetic effects in different groups, with the ensuing conditions i) FDR < 0.05 in one group AND *P* < 0.05 in another group, or ii) *P* _interaction_ < 0.05 AND *P* < 0.05 in both groups AND FDR _interaction_ < 0.005.

### Genetic association and mediation analyses

We ran a genome-wide association study (GWAS) in an European sample of 4,212 SLE patients and 4,065 controls previously described (28), and extracted the results for those SNPs involved in meQTLs regulating 148 DMPs. We established a Bonferroni significance threshold of *P* < 0.05/148 = 0.00033 to claim statistical significance. We performed mediation analyses with the R package mediation using the method development by Imai *et al*. (29) to quantify mediation effect of DNAm on SLE genetic risk. We report the Total Genetic Effect of the SNP on SLE (**c**), the Direct Genetic Effect of the SNP on SLE from a model adjusted by DNAm (**c’**), and estimated the proportion of the Total Genetic Effect on SLE explained by DNAm Mediation by the formula: *Prop. Mediated = (a*b)/(a*b+ c’)*, where ***a*** is the effect of the SNP on DNAm and ***b*** is the effect of DNAm in SLE. Test statistics for these measurements were estimated by 10,000 Monte Carlo simulations.

### Epigenetic associations with cytokine levels and transcription factor activities (TFact)

We inferred TFact for 119 Transcription Factors in the PRECISESADs data using whole blood RNAseq data and the R package DoRothEA (30) as previously described (31). Linear regression models were performed between DNAm at DMP and log transformed cytokine levels for 18 different inflammatory cytokines (see **supplementary Table 1**) or with TFact in different groups (SLE, CTRL, inflammatory and IFN) and corrected for the same covariates used for the EWAS/meQTLs. A Bonferroni threshold of *P* < 0.05/1,198 = 4E-5 was used to claim statistical significance.

### Identification of drug targets within epigenetic signals

We used different informatics platforms and data sources (**Supplementary File 1**) to identify drug targets within epigenetic signals. Specifically, for each gene in the list of the 549 differentially methylated genes, a “Total Score” was calculated as the sum of individual sub-scores, as described below (see also **Figure 6**):

**Figure 2.**
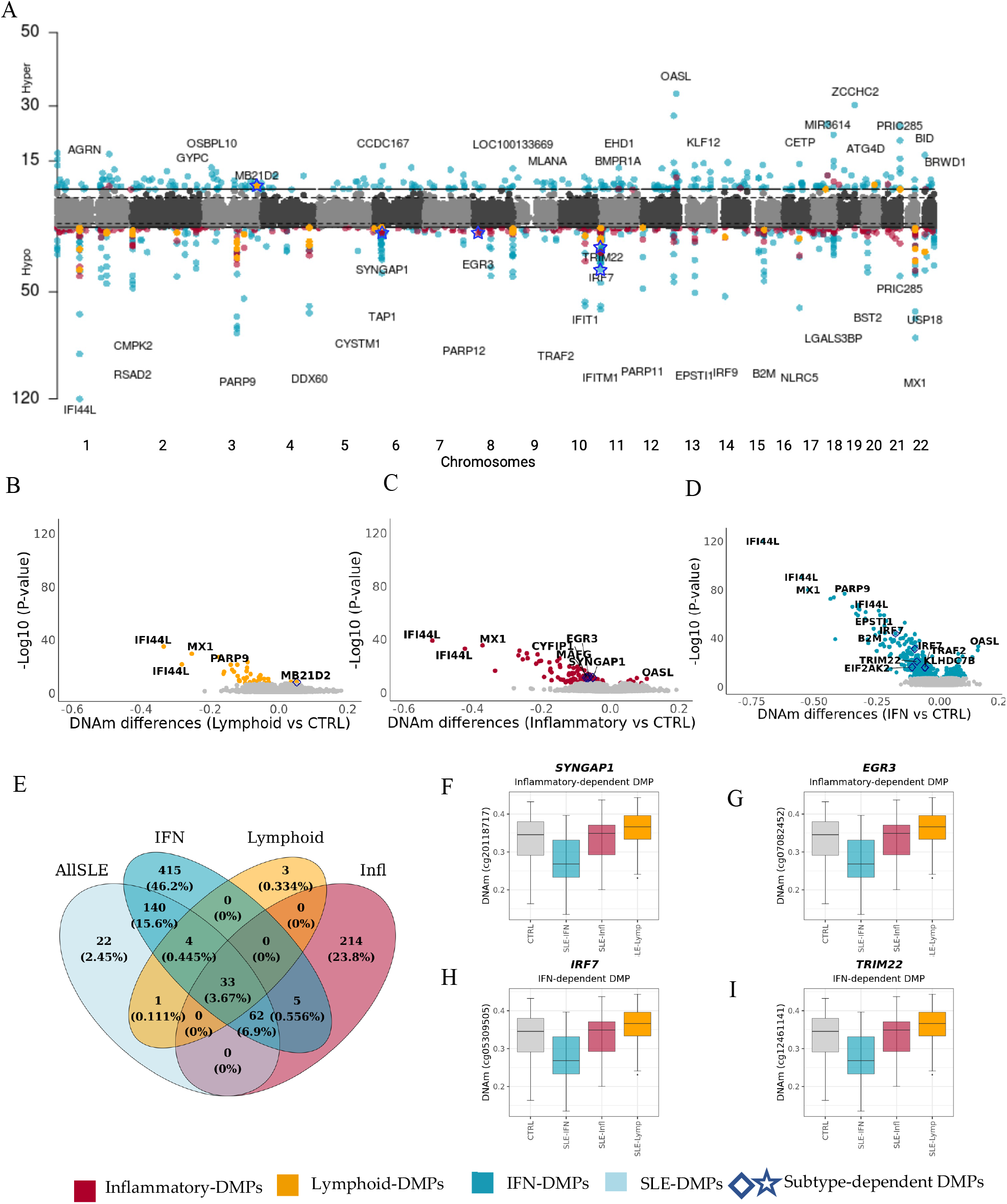
Epigenetic signatures of SLE molecular subtypes. **A**. Manhattan plot illustrating EWAS results for SLE and different molecular groups when compared with controls. X-axis represents the chromosomic locations of CpG sites and Y-axis represents the log10 (*P*) obtained in linear regression models. **B-D**. Volcano Plots representing EWAS results. The X-axis represents the DNAm differences between each pair of groups tested. **E**. Overlap of genome-wide significant results for each EWAS was performed. **F-I**. Examples of subtype dependent-DMPs. Colored dots represent significant DMPs after Bonferroni correction of different groups according to the legend. Diamonds and starts dots represents subtypes-dependent DMPs.

**Figure 3.**
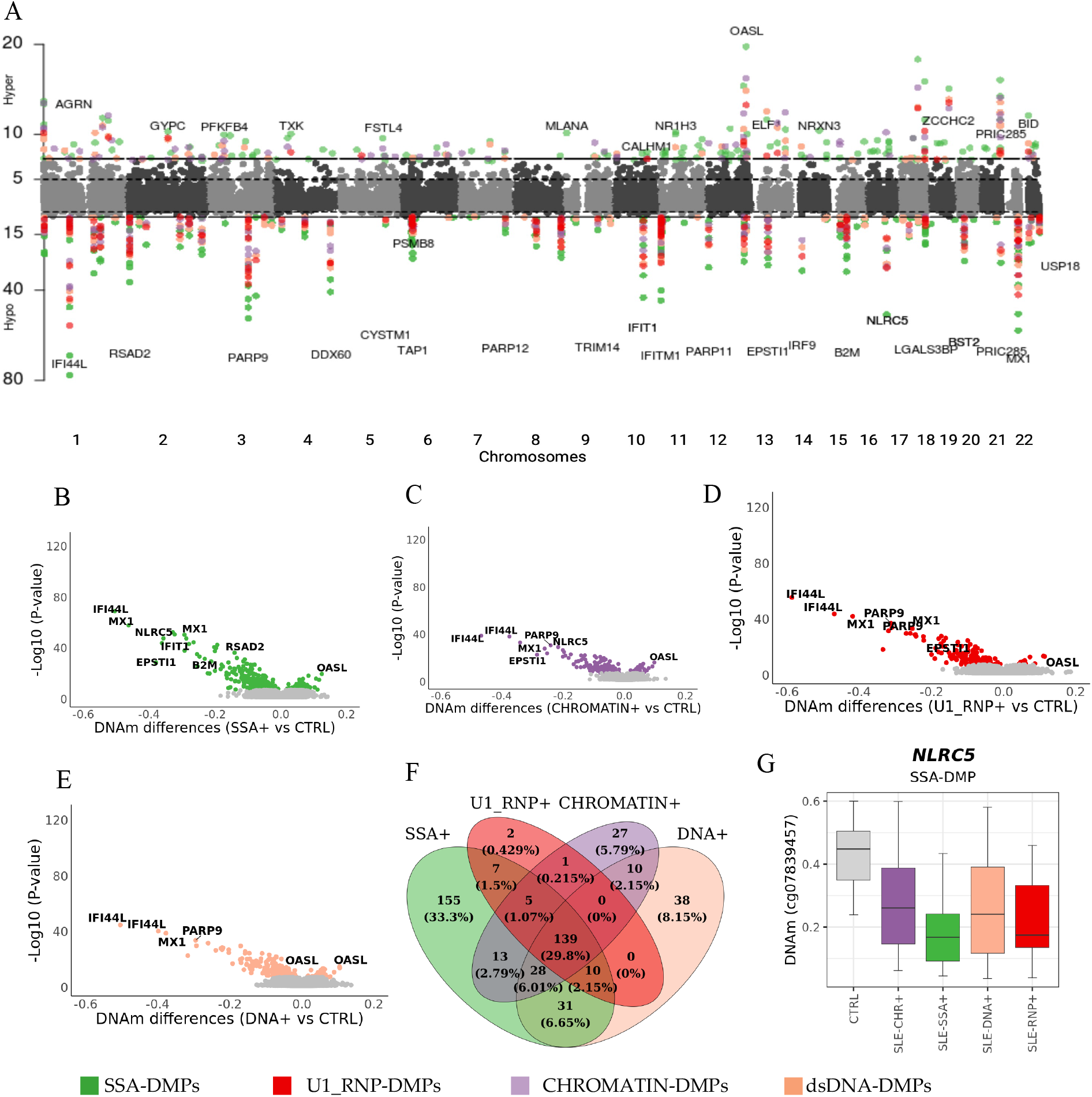
Epigenetic signatures of SLE autoantibody profiles. **A**. Manhattan plot illustrating EWAS results for different groups of SLE patients according to their autoantibody profiles when compared with controls. X-axis represents the chromosomal locations of CpG sites and Y-axis represents the log10 (*P*) obtained in linear regression models. **B-E**. Volcano Plots representing EWAS results. The X-axis represents the DNAm differences between each pair of groups tested. **F**. Overlap of genome-wide significant results for each EWAS performed. **G**. Example of DNAm distribution across different autoantibody groups, and healthy subjects. Colored dots represent significant DMPs after Bonferroni correction of different groups according to the legend.

**Figure 4.**
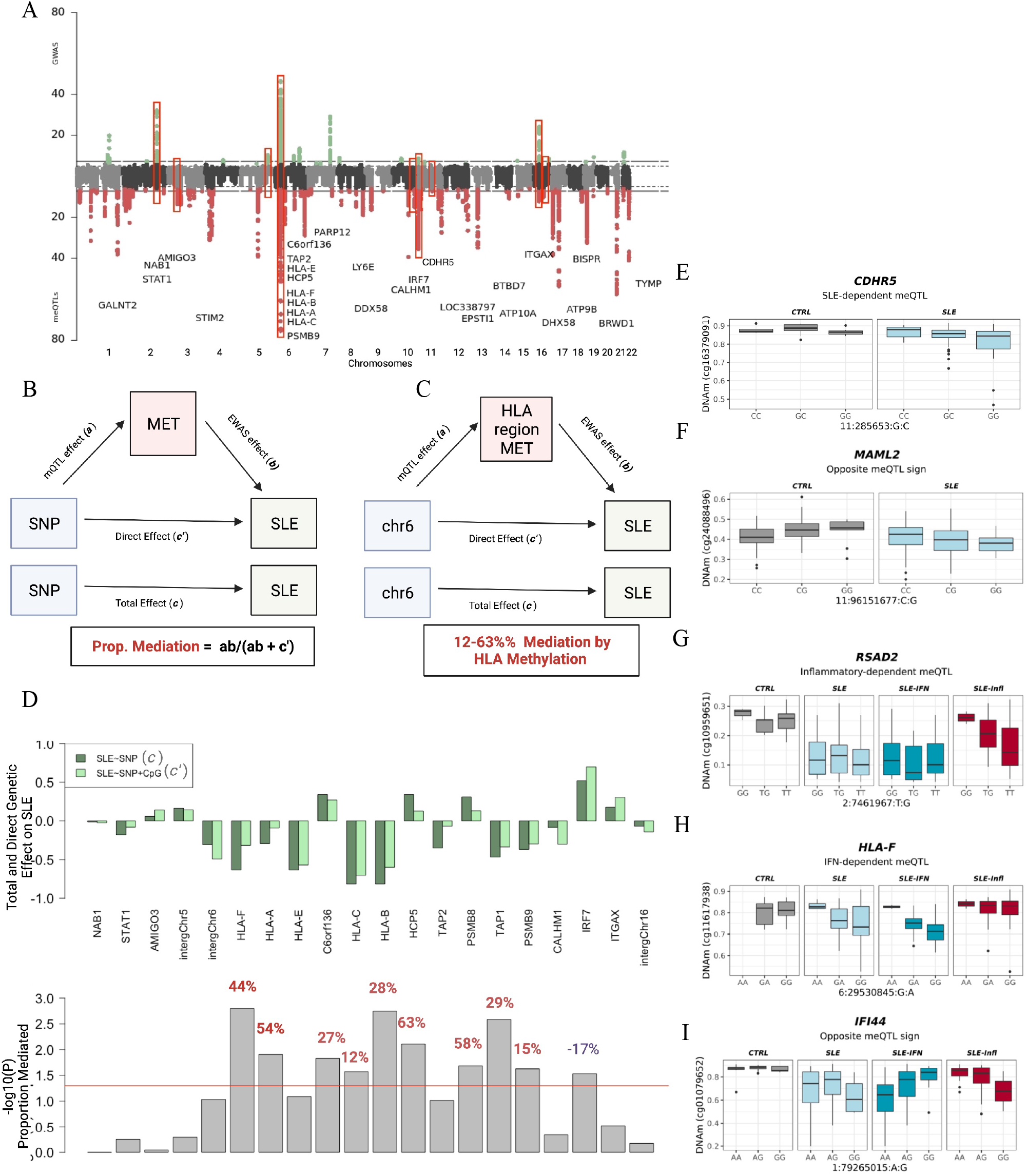
Genetic drivers of SLE-epigenetic signatures. **A**. Top Manhattan plot (MP) shows GWAS results contrasting allele frequencies between a group of SLE patients (N= 4,212) and a group of CTRL (N= 4,065). Bottom MP illustrates meQTL results for DMPs in the whole sample. X-axis represents the chromosomal locations of CpG sites and the Y-axis represents the log10 (*P*) obtained in a logistic regression model or meQTL analyses. Genetic associations above the red line marks the statistical association at a significant threshold of P < 1 × 10^−06^ for logistic associations and FDR < 0.05 for meQTLs. Red boxes show overlap in GWAS and meQTL results and represent meQTLs associated with SLE diagnosis. **B**. Mediation model in which SLE genetic risk is exerted partly through DNAm changes. **C**. Examples of SLE-associated SNPs in chr6 that are mediated by DNAm changes at DMPs in the HLA region. **D**. Mediation results for the best SLE-meQTL-DMPs by gene. Upper barplot shows the Total and the Direct Effect of SLE-associated genetic variants. Botton barplot show the significance of the proportion mediated via DNA met resulted from mediation models. Percentage of mediation is illustrated in red below each bar only for those significant genes (P _proportion mediated_ < 0.05) **E-J** Group-dependent meQTL effects. **E**. A meQTL significant effect is observed among SLE patients (FDR < 0.05) but not in the CTRL population (P>0.05). **F**. A meQTL significant effect is observed in SLE patients and CTRLs but with different signs. **G**. A meQTL significant effect is observed among SLE from inflammatory group (FDR < 0.05) but not in the CTRL population or in the IFN group (P>0.05). **H**. A meQTL significant effect is observed among SLE from IFN group (FDR < 0.05) but not in the CTRL population or in the inflammatory group (P>0.05). **I**. A meQTL with opposite direction effects between Inflammatory-IFN subtypes.

**Figure 5.**
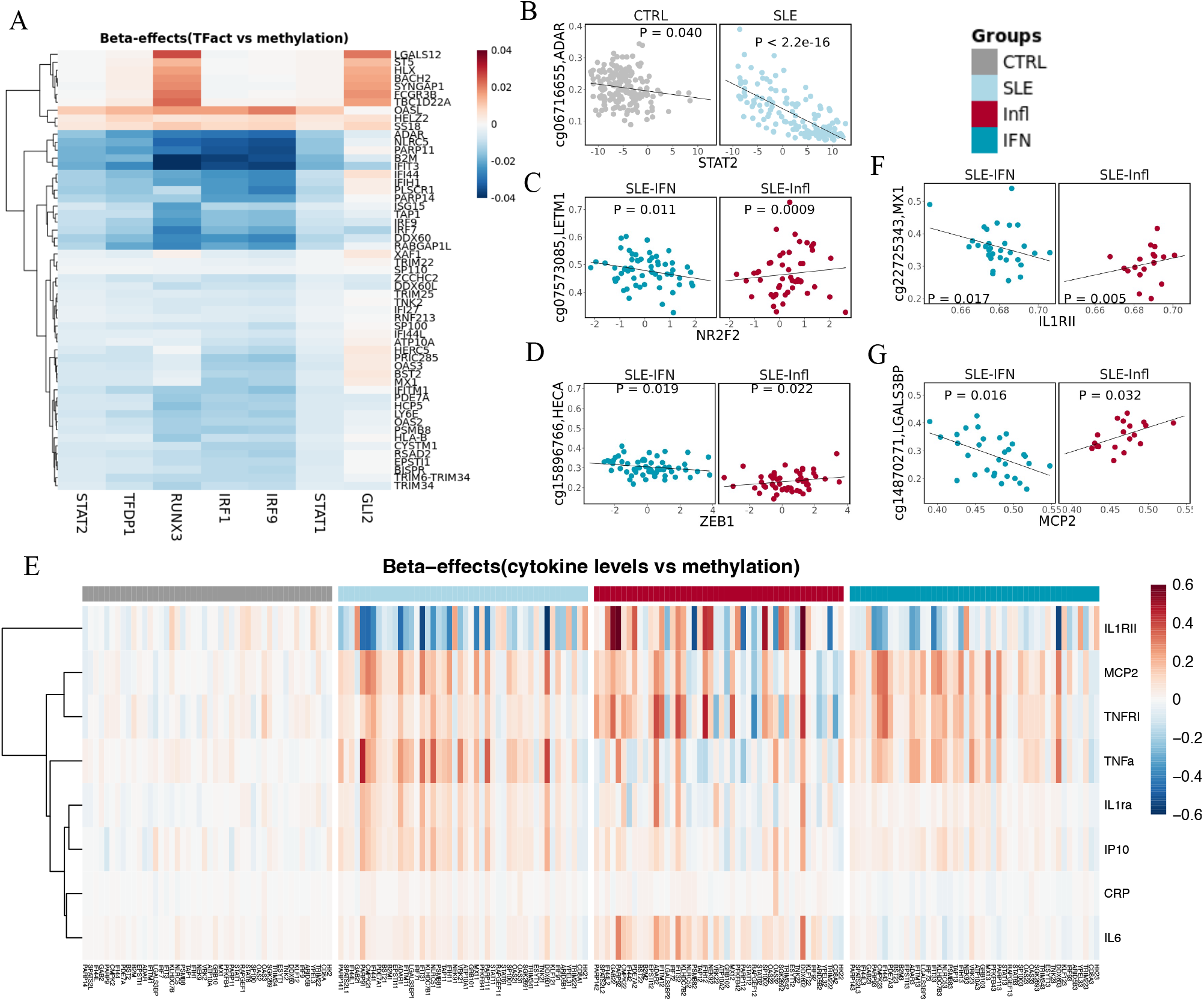
Relationship between SLE-epigenetic signatures, transcription factor activity, and cytokine production. **A**. Heatmap representing SLE-dependent associations between DNAm at DMPs and TFact inferred from RNAseq data. **B**. SLE dependent example showing the effect between TFact STAT2 and DNAm at *ADAR* gene vs CTRL. **C-D**. Subtypes dependent examples showing the effect between TFact *ZEB1* and *NR2F2* and DNAm at *SPATS2L* and *LETM1* genes. **E**. Heatmap showing the effect distribution of CpGs-genes associated to cytokine levels exhibiting group specificity. Color gradient from blue to red correspond to effect sizes. **F-G**. Examples for cytokine opposite associations at DNAm levels for inflammatory and IFN subtypes.

**Figure 6.**
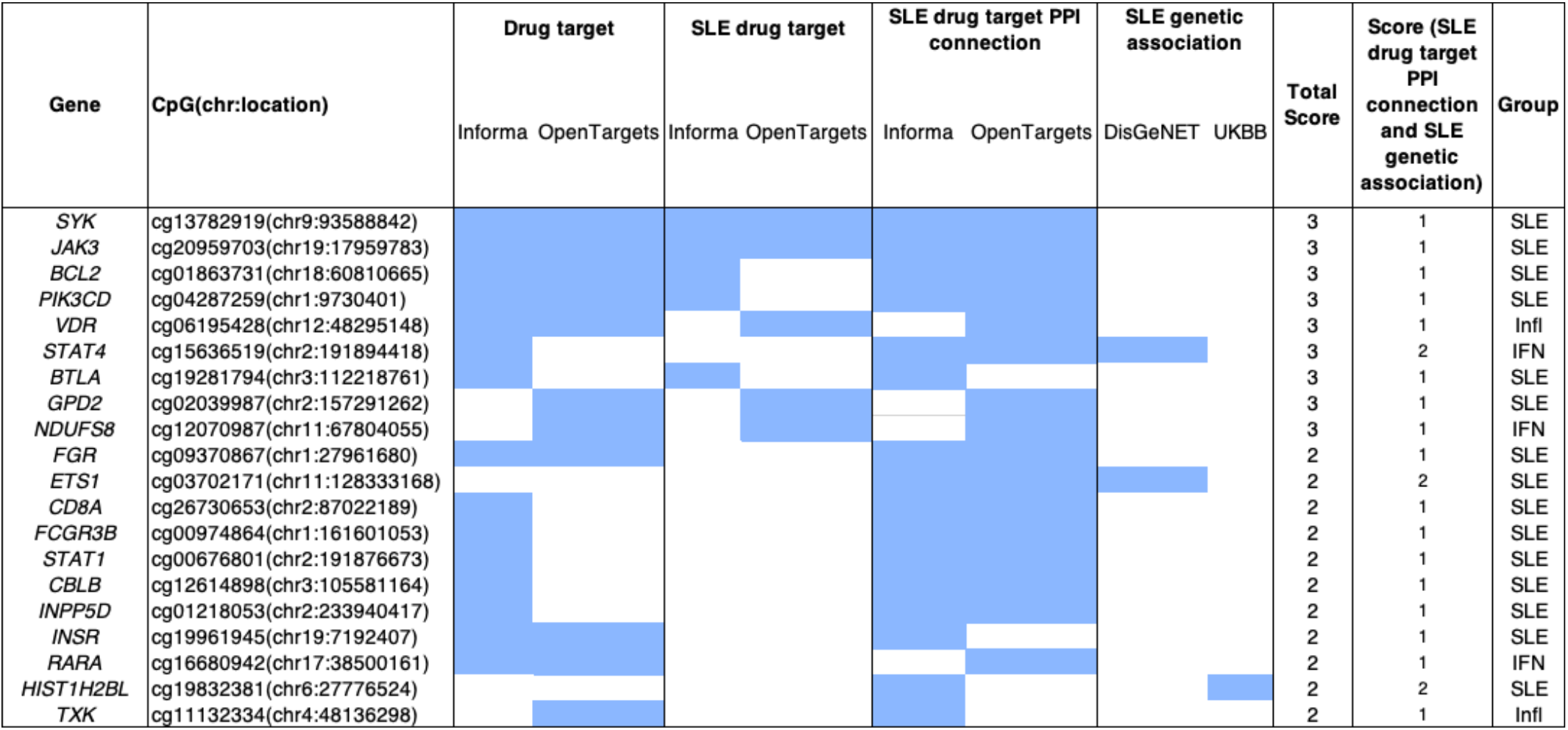
Representative SLE Epigenomic signature genes. We list the top 20 (of the 549 genes from the identified 974 CpG sites) based on summary of gene total scores derived from individual criteria (filled box indicates criterion satisfied). Filled boxes indicate an overlap with the data source described in each column. For full results, see **Supplementary Table 32**.

i. “known drug target” in OpenTargets OR Informa (clinical phase 1 or above), sub-score=1;
ii. “known drug target for SLE” in OpenTargets OR Informa (clinical phase 1 or above), sub-score=1
iii. having direct interactions with an SLE drug target (from OpenTargets OR Informa) in the PPI network, sub-score=1
iv. reported in DisGeNet to have GDA score of association with SLE > 0.3, sub-score=1
v. reported from UK BioBank with significant genetic variant associations (*P* < 5 × 10^−08^) to SLE, sub-score =1

## RESULTS

### 1. Genome-wide DNAm patterns associated with different molecular SLE subtypes

We identified 262 SLE-DMPs (97% replication rate in 450K independent sample) (**Figure 2A, Supplementary Table 2**), 64% of them exhibiting hypomethylation and 36% hypermethylation effects. Not unexpectedly, the top SLE-DMPs implicated large reductions of DNAm at IRGs and genes enriched in IFN pathways (**Supplementary Table 3**), as for example *IFI44L, MX1, NLRC5, IFITM1, IFIT1, IRF9* or *PARP9* but also genes involved in antigen processing and class I presentation (*TAP1* and *B2M*). Large hypermethylation effects were also observed in IRGs such as *OASL*. These results corroborate previous findings and is likely a consequence of the higher IFN levels observed in SLE patients (32).

Stratifying SLE patients into the molecular subtypes allowed us to raise the number of associations despite reducing the sample size (**Figure 2A**). We identified 314 inflammatory-DMPs (96.18% hypomethylation, **Supplementary Table 4**), 41 lymphoid-DMPs (90.24% hypomethylation, **Supplementary Table 5**) and 659 IFN-DMPs (54.48% hypomethylation, **Supplementary Table 6)**, among which 214, 3, and 420, respectively, had not been previously detected when all SLE patients were pooled together (**Figure 2E**). The IFN subtype exhibited the largest effects at genes involved in IFN pathways, but a persistent epigenIFNsig and similar enrichment were also observed in patients within the lymphoid and the inflammatory subtypes **(Figure 2B-D, Supplementary Table 3**). Inflammatory-DMG were also enriched in synaptic functions. (**Supplementary Table 7**).

We identified 71 inflammatory-dependent DMPs, 1 lymphoid-dependent DMPs and 232 IFN-dependent DMPs **(Supplementary Figure 1A-C, Supplementary Table 8)**. For example, we observed a DNAm decrease at *SYNGAP1* and *EGR3* genes in SLE inflammatory patients (*P* < 4.08 × 10^−04^) that we did not observe in healthy controls or in patients from other subtypes (*P* < 0.05) (**Figure 2F-G**). For the IFN subtype, we found large DNAm differences at *IRF7* and *TRIM22* genes, (**Figure 2H-I**) both of which are related to type I IFN pathways (33).

### 2. The relationship of autoantibody profile on the SLE epigenome

We stratified patients based on their positivity for the most prevalent AAbs. We identified 388 anti-SSA-DMPs, 223 anti-chromatin-DMPs, 256 anti-dsDNA-DMPs and 164 anti-U1-RNP-DMPs (**Figure 3A, Supplementary Tables 9-12**) when compared with CTRLs, yielding a total of 466 AAb-DMPs, from which 238 had not been previously detected as SLE-DMPs, and 81 were not detected as molecular subtype-DMPs. The epigenIFNsig, and in general every CpG effect, was stronger in SSA+ SLE patients for which we identified 155 DMPs not observed in the other groups (**Figure 3B-F**). However, the epigenetic signature was still persistent and dominant in SLE patients positive for other autoantibodies (**Figure 3B-E**). A small proportion of 27 anti-chromatin-DMPs and 38 anti-DNA-DMPs were exclusively observed in these groups when establishing a genome-wide significance level. However, at a significant threshold of P < 0.05, we could only identify one example of an anti-SSA-dependent-DMPs in the *NLCR5* gene that reached genome wide significance level in the SSA group and was not significant in the rest. (**Figure 3G, Supplementary Table 14**). Functional enrichment analyses on AAb-DMPs identified IFN pathways as well as antigen processing and other pathways regulatory of immune responses **(Supplementary Table 13)**. Our results indicate that epigenetic signature is highly shared across SLE patients with different autoantibody profiles, and that there is little specificity on SLE autoantibody-related epigenetic signals.

### 3. Genetic drivers of SLE epigenetic signals and mediation role of DNAm on SLE risk

We searched for *cis* genetic variants associated with DNAm at 148 DMPs (*cis*-meQTLs-DMPs, FDR < 0.05) (**Figure 4A, Supplementary Table 15**). Up to 31 loci involved in *cis*-meQTL-DMPs also associate with SLE (SLE-*cis*-meQTLs-DMPs, Bonferroni significance *P* < 8 × 10^−05^) (**Figure 4A, Supplementary Table 15)**. Mediation analyses on SLE-*cis*-meQTLs-DMPs (**Supplementary Table16)** revealed that SLE genetic risk is significantly reduced when DNAm is incorporated in the model (**Figure 4B**), and that a significantly high proportion of SLE genetic risk at the HLA class I region (12-65% proportion mediated) is mediated by DNAm in *HLA-F, HLA-A, C6orf136, HLA-C, HLA-B, HCP5, TAP1* and *PSMB9* genes (**Figure 4C-D)**. Other SLE-associated SNPs such as those located in *STAT1* or the intergenic region at chromosome 5 nearby microRNA mir-146 also showed a significant indirect effect of SNP on SLE suggesting a mediation role of DNAm, but the proportion mediated did not reach statistical significance (*P* > 0.05), probably due to the low sample size. Interestingly, for SLE-associated genes such as *IRF7* and *ITGAX*, the SLE genetic effect increased when DNAm was included in the model, suggesting a cofounded or more complex relationship between DNAm, genetic variation and SLE (**Figure 4D**).

### 4. Context-dependent meQTL regulatory function in SLE

We discovered *cis*-genetic variants associated with DNAm with dependency on disease status or molecular subtype by the identification of meQTL in different groups together with significant interaction effects **(Supplementary Table 17-19**). We identified SLE-dependent meQTLs for 394 DMPs (**Supplementary Table 20)**, among which significant disease-dependent effects were observed in *CDHR5* and *MAML2* **(Figure 4E-F**). Likewise, we discovered inflammatory- and IFN-dependent meQTLs for 283 and 316 DMPs, respectively **(Supplementary Table 21 and 22)**. For instance, we observed genetic regulation on DNAm in *RSAD2* gene in inflammatory SLE patients, but not in the IFN group or in controls where RSAD2-DNAm does not show a relationship with the genotype (**Figure 4G**). IFN-dependent meQTL effects were observed for example for *HLA-F* (**Figures 4H)**. Intriguingly, we also identified meQTLs with strong opposite effects (op-meQTLs) between SLE and CTRLs (119 DMPs) (**Supplementary Table 23**), or between the IFN and inflammatory subtypes (35 DMPs) among which *IFI44* gene shows the greatest opposite effect (**Figure 4I**) (**Supplementary Table 24**). Some genetic variants involved in context-dependent meQTLs were associated with SLE at a Bonferroni-corrected threshold (*P* < 1.2 × 10^−04^) (**Supplementary Table 17**). The strongest SLE-associated SNPs involved SLE-dependent meQTL that associated with DNAm at *HLA-B* and *HLA-E* genes. Interestingly, we also identified strong genetic-disease interaction (Pint = 6.5×10^−05^) in the regulation of DNAm within the *CDHR5* gene at chromosome 11, that involve SNPs strongly associated with SLE (P = 1.9×10^−07^) (**Figure 4E)**

### 5. SLE-associated epigenetic signals correlate with transcription factor activity and cytokine production

Transcription Factor (TF) binding has an important role in shaping DNAm levels and *vice versa* (34). Here, we identified up to 61 different TFs whose activity correlated with DNAm at several DMPs (P < 0.05) (**Supplementary Table 25**). Interestingly, the large interaction between DNAm and TFactivity for *IRF9, IRF1, STAT2, STAT1, STAT3, TFDP1, FOXM1, E2F3, E2F2, GLI2* and *RUNX3* was restricted to SLE patients (**Figure 5A-B, Supplementary Table 26**). While, TFs of IRF and STAT family have a well described role in the activation of IFN pathways and are associated with DNAm at IRGs. The TFact of *TFDP1, E2F3* and *FOXM1* correlated also with DNAm at IRGs such as *IFI44L, MX1, TRIM22* and *ISG15*. Interestingly, *RUNX3* activity was associated with DNAm at genes such as *FCGR3B, HLX, LGALS12, BACH* and *SYNGAP* which are not IFN-regulated. Intriguingly, we observed differential DNAm-TFact associations when comparing between inflammatory and IFN SLE patients (**Supplementary Table 27**). For example, *NR2F2* TFact is strongly negatively associated with DNAm at the *LETM1* gene, but such effect is not observed in the IFN subtype (**Figure 5C**). Likewise, *ZEB1* TF-act is associated with DNAm at *LGALS9, RSAD2, TMEM123, HECA* and *IFI44L* genes only in IFN patients (**Figure 5D**).

We explored whether DMPs could be associated with cytokine production in each molecular group (**Supplementary Tables 28-30**). In total, 82 DMPs were significantly associated with levels of 8 cytokines (P < 5 × 10^−05^). The strongest (and negative) associations were found between IL1RII levels and SLE-DMPs at a number of IRGs such as *PARP9* and *IFI44L*. We found SLE CpG-cytokine association for TNFa (*VRK2, PARP9*), IL1ra (*BST2, ATP10A*), IL1RII (*IFITM1, ARID5B*), MCP2 (*IFI44L, CMPK2*) and IP10 (*NLRC5, B2M*) which were not observed in CTRLs (**Supplementary Table 29**). We also observed inflammatory-dependent association for TNFRI (*RAPGEF1*) and CRP (*OAS3*) (**Supplementary Table 30**). **Figure 5E** illustrates the strength of the group-dependent associations. It can be observed that associations differed between groups and that for some CpGs, for cytokines such as IL1RII, these were stronger within the inflammatory as compared to the IFN subtype and stronger than CTRLs. For example, IL1RII was negatively associated with DNAm at *MX1* in IFN subtype but it is positively associated in the inflammatory subtype (**Figure 5F**), similarly MCP2 showed opposite effects at the DNAm level for the *LGALS3BP* gene for the inflammatory and IFN subtypes (**Figure 5G**). Altogether, our results indicate that the relationship between DNAm, TFact and cytokine production is determined in a subtype-dependent manner.

### 6. SLE-associated epigenetic signals inform drug discovery

We observed an enrichment of the list of unique 549 SLE differentially methylated genes (DMG) among known drug targets (Fold enrichment of 1.4, *P* < 0.01 in phase 1 or above in OpenTargets and Informa databases), and identified a total of 62 DMG being known drug targets, including 8 known SLE drug targets: *SYK, JAK3, BCL2, PIK3CD, VDR, BTLA, FGR, GDP2* and *NDUFS8* (**Supplementary Table 31-32, Figure 6**).

To identify and prioritize candidate novel SLE gene targets, we collected different features of each gene and ranked them based on two criteria: i) previous evidence of genetic association to SLE ii) “network proximity” (**Figure 6**, see **Methods** for scoring details). We showed strong enrichment of validated targets within the groups of genes with scores equal or higher than one (**Supplementary Figure 2**). Using the different annotations available for each gene of the list, we could prioritize genes of interest and focus on potentially novel targets (see **discussion**) for SLE, and for different SLE subtypes. For example, we observed that 4 of the known drug targets (*VDR, ALOX5AP, ITGA5* and *ECE1*) overlapped with genes that are differentially methylated only in the inflammatory SLE patients. Likewise, 17 genes (*CLU, CETP, TSHR, ITGA2, ACACA, TNK2, STAT4, PSMB8, SCN8A, INPP5D, BMPR1A, TAP1, TYMP, QPCT, GPD2, PSMB9* and *LAMB1*) overlapped with interferon specific epigenetic signatures (**Supplementary Table 32**).

## DISCUSSION

In this study we present the first integrative EWAS that contrasts genome-wide DNAm data in several stratifications of SLE patients and integrates results with genetic, transcription, clinical and serological data.

Stratifying SLE patients across homogenous molecular subtypes allowed us to significantly increase statistical power expanding the epigenetic signals and reporting new loci associated with SLE that had not been revealed when analyzing SLE heterogeneous populations. The largest effects were mostly found at interferon related genes as for example *IFI44L, MX1, NLRC5, PARP9*/*DTX3L*, which has been extensively identified in previous SLE-EWAS across most blood cell types and in other SADs(7). Here we show that the epigenIFN sig is present across all SLE molecular subtypes at different intensities, however we could discover some epigenetic signals exclusively in those SLE patients from the IFN subtype, as DMPs at *IRF7* and *TRIM22* genes. *IRF7* has been established as an SLE genetic risk locus that alters IFN type I expression (33) and TRIM molecules have been studied as autoantigens in some autoimmune diseases, especially in Sjögren’s syndrome (SS) where *TRIM22* protein showed little or no immunoreactivity in a sub-population of SS patients(35) suggesting that *TRIM22* was acting as a potential SSA60 regulated gene. Importantly, we also discovered epigenetic signals that are specific of SLE patients with an inflammatory molecular profile, as those in the *EGR3* and *SYNGAP* genes. *EGR3* is a member of a zinc finger transcription factor that plays an important role in regulating immune responses inducing also the expression of anti-inflammatory cytokines such as IL-10 and TGFB1 (31), however the role of EGR3 in autoimmunity it is not clear yet. *SYNGAP1* encodes a Ras GTPase activating protein that is member of the N-methyl-D-aspartate receptor complex and has been identified as a differentially methylated gene associated to gene expression in systemic sclerosis (SSc)(36).

Our stratification approach also included an exploration of the AAb relationship on the SLE epigenome. The DMPs with largest effects were found within SSA positive SLE patients. Our previous work showed that the IFNepig signature in Sjogren’s syndrome is restricted to SSA positive patients and driven by HLA genetic variation(9). However, in SLE many of the signals observed are shared across SLE patients with different AAb profiles, which suggests that in SLE, the epigenetic signature is not AAb-specific or AAb specificities are highly correlated.

One of the most important findings of this work is that SLE-associated genetic variation might exhibit its risk through DNAm changes, this is especially true for SLE-associated SNPs within the HLA region. Our findings strongly suggest that beyond the impact on antigen presentation, genetic risk at the HLA region is also mediated through the epigenetic and transcriptional alterations of many genes residing within the HLA such as *HLA-F, HLA-A, HLA-C, HCP5, PSMB8, TAP1* and *PSMB9*. These results are in concordance with previous work showing an overexpression of these genes in SLE patients (7). Outside the HLA region, our mediation results also support an indirect effect of SLE-associated genetic variation via DNAm within *STAT1* and *microRNA146* gene, previously genetically associated with SLE. However, given the small effects of these SNPs on SLE, and the sample size analyzed here, confirmatory studies are needed to provide conclusive results. In the same line, analyzing public repositories of genetic associations by means of Mendelian Randomization approaches would help to resolve puzzling relationships as those observed for genetic variation, DNAm and SLE risk at the *IRF7* gene.

Our findings show that there exists a clear influence of disease and molecular status on the genetic architecture of DNAm, given the fact that DNAm at a large proportion of DMPs is regulated by meQTLs but exclusively in the SLE population context or in certain molecular states. For example, this is the case for *CDHR5* gene which DNAm is regulated by genetics only in the IFN and SLE subtypes, respectively. Interestingly, our results show that genetic variants involved in CDHR5 meQTLs are also associated with SLE risk. The SLE-associated SNPs represent novel disease variants as they have not been previously identified in GWAS and are likely to mediate their effects trough DNAm changes. Previous studies have identified *CDHR5-*SNPs located close to *IRF7* (37,38) and associated with systemic sclerosis (SS) (38). The candidate gene *CDHR5* is a member of the cadherin family which interacts with the β-catenin pathway (39). These observations are in agreement with agrowing body of evidence that highlights that genetic effects are largely context and time-specific, and implicates that future research and data collection of pathological and molecular status within longitudinal larger autoimmune populations will be able to decipher many more genetic variants with important regulatory functions in autoimmunity.

TF activity (TFact) has been linked to SLE recently (31). In this work, we identified a group of TFs whose activity was associated with SLE-associated DNAm changes. The activity of STAT and IRFs regulators show the strongest association with DMPs at IRGs, and here we show that this relationship is specific of SLE population and not observed in healthy individuals. We also reveal other TFs not previously implicated in SLE, such as *TFDP1, E2F3* and *FOXM1* whose activity associates with DNAm at IRGs. On the contrary, we identified that *RUNX3* activity, a susceptibility gene in SLE and systemic sclerosis (SSc) (40,41), correlates with DNAm at non-IFN related genes. Our results suggest that TFact could play a relevant role in autoimmunity by altering epigenetic programs.

In SLE pathogenesis, an inflammatory cascade is mediated by altered cytokine production. Despite the growing recognition of the high potential of DNAm changes as surrogate and biomarkers of pro-inflammatory proteins, especially in ageing phenotypes (42,43), no study, to the best of our knowledge, had reported epigenetic-cytokine correlations within autoimmune diseases. Here we show a strong association in the SLE population between inflammatory cytokines and methylation changes, not observed within healthy individuals, being IL2RA and IP10 the cytokines showing the strongest association with the epigIFNsign. Our results also show that many epigenetic-cytokine associations are only observed in certain molecular contexts.

Finally, we discovered several interesting potential new drug targets based on the SLE epigenetic profiles. Among the novel SLE candidate genes with higher score is *STAT4*, which is in direct PPI connections with SLE drug targets and exhibit strong genetic associations (44–49). Another interesting target candidate is the transcription Factor *ETS1*, which also had a significant strong SLE genetic association (50) and which is in direct PP interaction with SLE drug targets such as *JAK3*. This protein has been observed to suppress T follicular helper type 2 cell differentiation and halt SLE onset (51,52). Of interest, FGR, a member of the Src family of protein tyrosine kinases (PTKs) was scoring high for being a drug target for non-SLE conditions, but in close PPI connections with SLE validated targets, making it a potential candidate for drug repositioning. It functions as a positive regulator of cell migration and regulates cytoskeleton reorganization via RAC1 activation. It also phosphorylates SYK and promotes SYK-dependent activation of AKT1 and MAP kinase signaling. *SYK* is one of the top genes in the candidate drug target gene list for SLE (31,53), as *STAT4* is for the IFN subytpe and *VDR* for the inflammatory.

To wrap up, our study disentangles epigenetic signatures in SLE with regards to different heterogeneity aspects and identifies potential novel drug targets. By means of integrative multi-omics analyses we show that the strong association between epigenetics and genetics, TF activity and cytokine production is highly dependent on disease and molecular context.

## Data Availability

The cohort datasets generated and analyzed during the current study are available upon request through ELIXIR platform.

## Acknowledgements

E.C.M and O.C.P have been funded by Grants for the financing of Research, Development and Innovation (R + D + I) in Biomedicine and Health Sciences in Andalusia for the year 2021, project co-financed by 80% by funds of the FEDER Operational Program of Andalusia 2014-2020 (File Number: PECOVID-0072-2020).

Funding for the preparation of this manuscript has received support from the Innovative Medicines Initiative Joint Undertaking under grant agreement no 115,565, resources composed of the financial contribution from the European Union’s Seventh Framework Program (FP7/2007-2013) and the EFPIA companies’ in kind contribution.

Funding for this work has been provided by the Lupus Research Alliance/BMS Award to M.E.A.R. and the Diversity Supplement to O.C.P.

This project has received funding from the Innovative Medicines Initiative 2 Joint Undertaking (JU) under grant agreement No 831434 (3TR). The JU receives support from the European Union’s Horizon 2020 research and innovation programme and EFPIA.

## Ethics declarations

An ethical protocol was prepared, reached consensus across all partners, academic and industrial, translated into all participant’s languages and approved by each of the local ethical committees of the clinical recruitment centers, and all experimental protocols were approved by each of the local committees. For a list of local committees and centers involved in PRECISESADS please see Supplementary Note <u>1</u>. All patients recruited to the study were aged 18 years or older and signed an informed consent form, and all methods were carried out in accordance with relevant guidelines and regulations. The study adhered to the standards set by International Conference on Harmonization and Good Clinical Practice, and to the ethical principles that have their origin in the Declaration of Helsinki (2013). The protection of the confidentiality of records that could identify the included individuals is ensured as defined by the EU Directive 2001/20/EC and the applicable national and international requirements relating to data protection in each participating country.

## Author contributions

E.C.-M., M.E.A.-R., E.dR. and O.C.P contributed to the conception and design of the study and also drafted the manuscript. The PRECISESADS Clinical Consortium members agreed on the clinical data, recruited the patients, and performed their detailed clinical assessment. PRECISESADS Cytometry Consortium obtained flow cytometry data. O.C.P, E.C.-M., G.B., M.K., R.L.T, D.R., A.I., E.dR., W.Q., C.Z., S.S., A.S. and M.M.-B. contributed to pre-processing and/ or analyzing the data. M.E.A.-R., J.M., C.M., E.dR and E.B. contributed to data generation and sample recruitment. M.E.A.-R. coordinated the entire PRECISESADS project. All authors read and approved the final manuscript.

## Competing interests

Weiliang Qiu, Cheng Zhu and Srinivas Shankara are employees of SANOFI. All other authors have no competing interests.

**PRECISESADS Clinical Consortium**

**Lorenzo Beretta**^**9**^, **Barbara Vigone**^**9**^, **Jacques-Olivier Pers**^**10**^, **Alain Saraux**^**10**^, **Valérie Devauchelle-Pensec**^**10**^, **Divi Cornec**^**10**^, **Sandrine Jousse-Joulin**^**10**^, **Bernard Lauwerys**^**11**^, **Julie Ducreux**^**11**^, **Anne-Lise Maudoux**^**11**^, **Carlos Vasconcelos**^**12**^, **Ana Tavares**^**12**^, **Esmeralda Neves**^**12**^, **Raquel Faria**^**12**^, **Mariana Brandão**^**12**^, **Ana Campar**^**12**^, **António Marinho**^**12**^, **Fátima Farinha**^**12**^, **Isabel Almeida**^**12**^, **Miguel Angel Gonzalez-Gay Mantecón**^**13**^, **Ricardo Blanco Alonso**^**13**^, **Alfonso Corrales MartÍnez**^**13**^, **Ricard Cervera**^**14**^, **Ignasi RodrÍguez-Pintó**^**14**^, **Gerard Espinosa**^**14**^, **Rik Lories**^**15**^, **Ellen De Langhe**^**15**^, **Nicolas Hunzelmann**^**16**^, **Doreen Belz**^**16**^, **Torsten Witte**^**17**^, **Niklas Baerlecken**^**17**^, **Georg Stummvoll**^**18**^, **Michael Zauner**^**18**^, **Michaela Lehner**^**18**^, **Eduardo Collantes**^**19**^, **Rafaela Ortega Castro**^**19**^, **Ma Angeles Aguirre-Zamorano**^**19**^, **Alejandro Escudero-Contreras**^**19**^, **Ma Carmen Castro-Villegas**^**20**^, **Norberto Ortego**^**20**^, **MarÍa Concepción Fernández Roldán**^**20**^, **Enrique Raya**^**21**^, **Inmaculada Jiménez Moleón**^**21**^, **Enrique de Ramon**^**22**^, **Isabel DÍaz Quintero**^**22**^, **Pier Luigi Meroni**^**23**^, **Maria Gerosa**^**23**^, **Tommaso Schioppo**^**23**^, **Carolina Artusi**^**23**^, **Carlo Chizzolini**^**24**^, **Aleksandra Zuber**^**24**^, **Donatienne Wynar**^**24**^, **Laszló Kovács**^**25**^, **Attila Balog**^**25**^, **Magdolna Deák**^**25**^, **Márta Bocskai**^**25**^, **Sonja Dulic**^**25**^, **Gabriella Kádár**^**25**^, **Falk Hiepe**^**26**^, **Velia Gerl**^**26**^, **Silvia Thiel**^**26**^, **Manuel Rodriguez Maresca**^**27**^, **Antonio López-Berrio**^**27**^, **RocÍo Aguilar-Quesada**^**27**^ **& Héctor Navarro-Linares**^**27**^

^9^Referral Center for Systemic Autoimmune Diseases, Fondazione IRCCS Ca’ Granda Ospedale Maggiore Policlinico di Milano, Milano, Italy. ^10^Centre Hospitalier Universitaire de Brest, Hospital de la Cavale Blanche, Brest, France. ^11^Pôle de pathologies rhumatismales systémiques et inflammatoires, Institut de Recherche Expérimentale et Clinique, Université catholique de Louvain, Brussels, Belgium. ^12^Centro Hospitalar do Porto, Porto, Portugal. ^13^Servicio Cantabro de Salud, Hospital Universitario Marqués de Valdecilla, Santander, Spain. ^14^Hospital Clinic I Provicia, Institut d’Investigacions Biomèdiques August Pi i Sunyer, Barcelona, Spain. ^15^Katholieke Universiteit Leuven, Leuven, Belgium. ^16^Klinikum der Universitaet zu Koeln, Cologne, Germany. ^17^Medizinische Hochschule Hannover, Hannover, Germany. ^18^Medical University Vienna, Vienna, Austria. ^19^Servicio Andaluz de Salud, Hospital Universitario Reina SofÍa Córdoba, Córdoba, Spain. ^20^Servicio Andaluz de Salud, Complejo hospitalario Universitario de Granada (Hospital Universitario San Cecilio), Granada, Spain. ^21^Servicio Andaluz de Salud, Complejo hospitalario Universitario de Granada (Hospital Virgen de las Nieves), Granada, Spain. ^22^Servicio Andaluz de Salud, Hospital Regional Universitario de Málaga, Málaga, Spain. ^23^Università degli studi di Milano, Milan, Italy. ^24^Hospitaux Universitaires de Genève, Genève, Switzerland. ^25^University of Szeged, Szeged, Hungary. ^26^Charite, Berlin, Germany. ^27^Andalusian Public Health System Biobank, Granada, Spain.

**PRECISESADS Flow Cytometry Study Group**

**Montserrat Alvarez**^**28**^, **Damiana Alvarez-Errico**^**29**^, **Nancy Azevedo**^**30**^, **Nuria Barbarroja**^**31**,**32**^, **Anne Buttgereit**^**33**^, **Qingyu Cheng**^**34**^, **Carlo Chizzolini**^**28**^, **Jonathan Cremer**^**35**^, **Aurélie De Groof**^**36**^, **Ellen De Langhe**^**37**^, **Julie Ducreux**^**36**^, **Aleksandra Dufour**^**28**^, **Velia Gerl**^**34**^, **Maria Hernandez-Fuentes**^**38**^, **Laleh Khodadadi**^**34**^, **Katja Kniesch**^**39**^, **Tianlu Li**^**34**^, **Chary Lopez-Pedrera**^**36**^, **Zuzanna Makowska**^**33**^, **Concepción Marañón**^**41**^, **Brian Muchmore**^**41**^, **Esmeralda Neves**^**30**^, **Bénédicte Rouvière**^**42**^, **Quentin Simon**^**42**^, **Elena Trombetta**^**40**^, **NievesVarela**^**39**^ **&TorstenWitte**^**39**^

^28^Immunology and Allergy, University Hospital and School of Medicine, Geneva, Switzerland. ^29^Chromatin and Disease Group, Bellvitge Biomedical Research Institute (IDIBELL), Barcelona, Spain. ^30^Serviço de Imunologia EX-CICAP, Centro Hospitalar e Universitário do Porto, Porto, Portugal. ^31^IMIBIC, Reina Sofia Hospital, University of Cordoba, Córdoba, Spain. ^32^Bayer AG, Berlin, Germany. ^33^Pharmaceuticals Division, Bayer Pharma, Berlin, Germany. ^34^Department of Rheumatology and Clinical Immunology, Charité University Hospital, Berlin, Germany. ^35^Department of Microbiology and Immunology, Laboratory of Clinical Immunology, KU Leuven, Leuven, Belgium. ^36^Pôle de Pathologies Rhumatismales Inflammatoires et Systémiques, Institut de Recherche Expérimentale et Clinique, Université Catholique de Louvain, Brussels, Belgium. ^37^University Hospitals Leuven and Skeletal Biology and Engineering Research Center, KU Leuven, Leuven, Belgium. ^38^UCB, Slough, UK. ^39^Klinik für Immunologie Und Rheumatologie, Medical University Hannover, Hannover, Germany. ^40^Laboratorio di Analisi Chimico Cliniche e Microbiologia - Servizio di Citofluorimetria, Fondazione IRCCS Ca’ Granda Ospedale Maggiore Policlinico di Milano, Milan, Italy. ^41^GENYO, Center for Genomics and Oncological Research Pfizer/University of Granada/Andalusian Regional Government, Granada, Spain. ^42^NSERM, UMR1227, CHRU Morvan, Lymphocytes B et Autoimmunité, University of Brest, BP 824, Brest, France.

## FIGURE LEGENDS

**Supplementary Figure 1. Effect Size Comparison comparing results from different EWAS. A**. Effect size comparison between inflammatory-dependent DMPs and lymphoid-dependent DMPs. **B**. Effect size comparison between inflammatory-dependent DMPs and IFN-dependent DMPs. **C**. Effect size comparison between IFN-dependent DMPs and lymphoid-dependent DMPs.

**Supplementary Figure 2. Relationship between SLE Epigenomic signature genes and known drug targets. A**. Bar plot with number of methylated genes and known drug targets that are in these methylated genes under different score thresholds. **B**. Bar plot with number of methylated genes and the rate of known drug targets in these methylated genes under different score thresholds.

