## Supplementary File 1 for "The Lupus Epigenome Relates to Genetics, Transcription and Serological Profiles with Dependency on Molecular Subtypes and Informs Drug Discovery"

Details of platforms and date sources used for scores of methylated genes

1. *The Open Targets (OT) platform* (https://www.opentargets.org/) is a public-private research partnership aimed at integration of multiple target-disease linkage evidence and the development of exploratory methods to facilitate drug target selection and validation. A broad range of target-disease association features were aggregated in OT from public domain information sources, specifically genetic association, somatic mutation, pathway biology, transcriptomics, text mining, animal model, and known drug target status.
2. *Informa* is one of the largest databases for global clinical trials. It aggregates drug, drug targets and clinical trial information from over 40,000 data sources in the public domain, including company press releases, government drug and trial databases (e.g., Drugs@FDA and ClinicalTrials.gov), and scientific conferences and publications. The full dataset can be accessed online at https://citeline.informa.com/.
3. *DisGeNET* (http://www.disgenet.org) consists of large collections of gene to human disease and phenotype associations. The database collects the information from multiple sources such as expert curated repositories, GWAS catalogues, animal models and scientific literature. A gene-disease association (GDA) score (ranging from 0 to 1) is reported as an estimate of the strength of each association.
4. *UK Biobank(UKBB).* Genetic associations were retrieved from GWAS summary statistics from UKBB (https://www.ukbiobank.ac.uk/). A p-value threshold of 5 × 10-8 was used to identify significant genetic variant associations to SLE trait.
5. *PPI network* was from STRING database (https://string-db.org/, version11). The STRING database is one of the most comprehensive protein to protein interaction network with predicted and known interactions. Each edge is given a weight to identify the degree of confidence. In order to generate a reliable, high-trust level network reference in this work, we selected interactions with confidence score greater than 0.7 defined by STRING. After data preprocessing, we reconstructed our global protein-protein interaction network with 16,795 nodes and 252,013 edges.
