## Supplementary Figure 1 for "The Lupus Epigenome Relates to Genetics, Transcription and Serological Profiles with Dependency on Molecular Subtypes and Informs Drug Discovery"

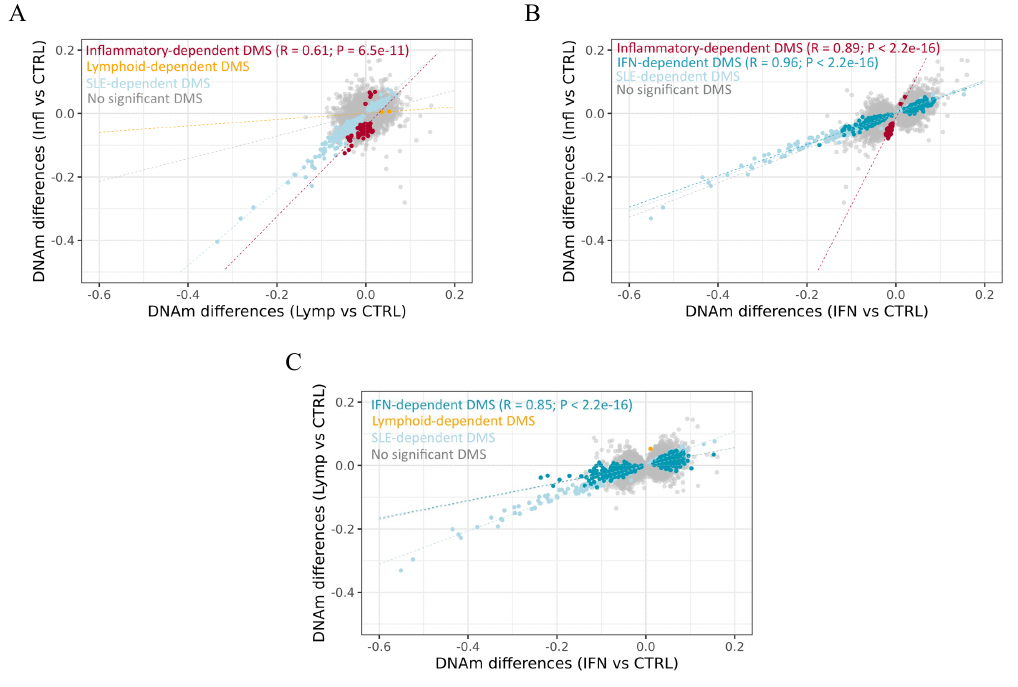
**Supplementary Figure 1.** **Effect Size Comparison comparing results from different EWAS**. **A**. Effect size comparison between inflammatory-dependent DMS and lymphoid-dependent DMS. **B**. Effect size comparison between inflammatory-dependent DMS and IFN-dependent DMS. **C**. Effect size comparison between IFN-dependent DMS and lymphoid-dependent DMS.
