## Supplementary Figure 2 for "The Lupus Epigenome Relates to Genetics, Transcription and Serological Profiles with Dependency on Molecular Subtypes and Informs Drug Discovery"

**Supplementary Figure 2.** Relationship between SLE Epigenomic signature genes and known drug targets. We set different score thresholds to reduce the number of SLE methylated genes and compare them with known drug targets. The score threshold was set based on the portion of total scores that derived from PPI direct interactions with an SLE drug target and SLE genetics association analysis (Figure 6). **A.** Bar plot with number of methylated genes and known drug targets that are in these methylated genes under different score thresholds. **B.** Bar plot with number of methylated genes and the rate of known drug targets in these methylated genes under different score thresholds.

**A.**

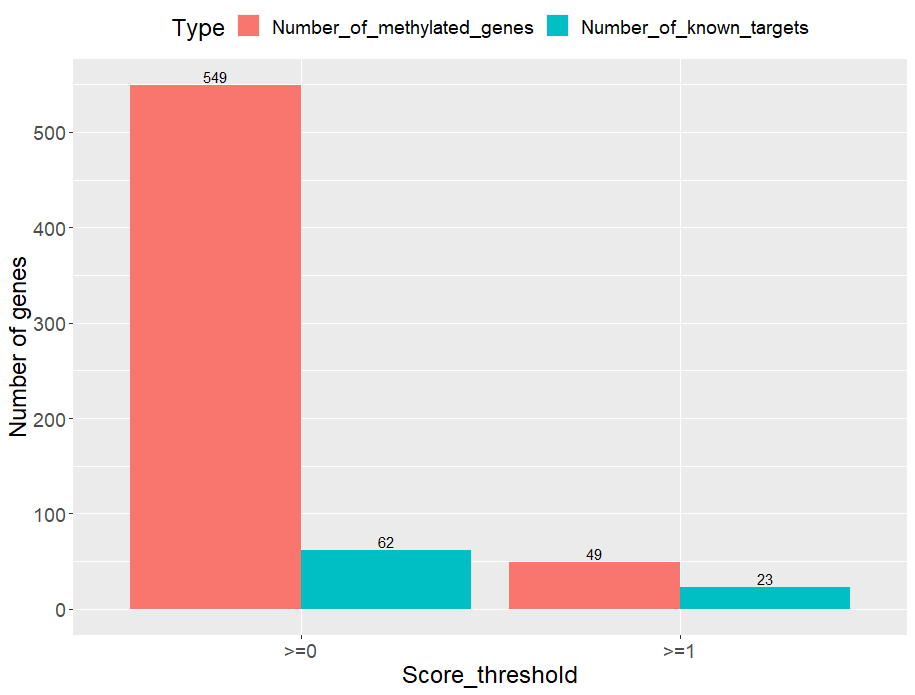

**B.**

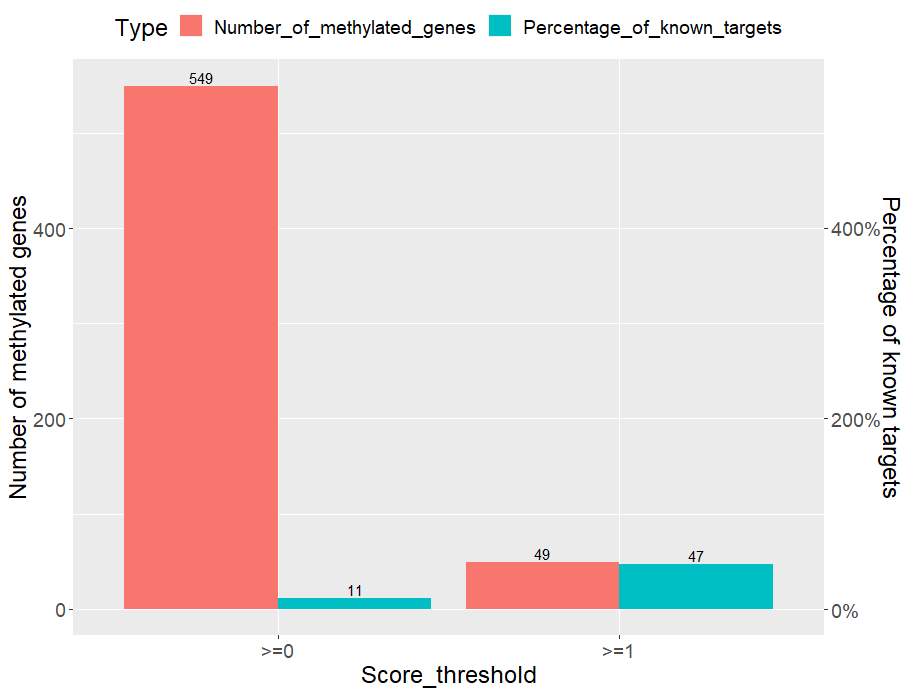
